# White Matter Hyperintensities Impact Biomarker Prediction of Cognitive Decline

**DOI:** 10.1101/2025.08.09.25333299

**Authors:** Dario Bachmann, Christoph Gericke, Maha Wybitul, Antje Saake, Sandro Studer, Katrin Rauen, Esmeralda Gruber, Andreas Buchmann, Martin Hüllner, Kaj Blennow, Henrik Zetterberg, Roger M Nitsch, Christoph Hock, Valerie Treyer, Anton Gietl

## Abstract

**Introduction:** Accounting for common co-pathologies such as white matter hyperintensities (WMH) of presumed vascular origin may improve the prognostic performance of imaging and blood-based biomarkers for cognitive decline in Alzheimer’s disease (AD).

**Methods:** We analyzed longitudinal cognitive data from 216 (median age: 67.2 years; median follow up: 5.3 years) community-dwelling older adults who underwent MRI, Aβ PET, and plasma p-tau217, neurofilament light chain, and glial fibrillary acidic protein assessment.

**Results:** WMHs predicted declines in executive function, processing speed, and global cognition, and amplified the effects of imaging and plasma biomarkers, especially on episodic memory. The interaction between Aβ-PET and WMH volume was the primary interaction associated with faster memory decline.

**Discussion:** Coexisting AD pathology and WMHs shorten the preclinical stage of AD. Consideration of Aβ-PET and blood-based biomarkers in the context of an individual’s WMH burden may improve the prognostic value of these biomarkers.

## 1. Background

Accurate prediction of cognitive decline and clinical progression in preclinical and early Alzheimer’s disease (AD) is important for personalized prognosis and better patient stratification in clinical trials. An important tool for determining who is likely to suffer from cognitive decline is amyloid-β (Aβ) positron emission tomography (PET) imaging.^1^ More recently, blood-based biomarkers have shown great potential in predicting disease progression in both early and late stages of AD.^2^ These blood biomarkers offer significant advantages in terms of accessibility and cost-effectiveness compared to PET imaging. Despite these advancements in biomarker-based prognosis of disease progression, predicting the speed of cognitive decline in AD remains challenging because disease courses are highly heterogeneous. One factor that contributes to individual differences in cognitive trajectories is the presence of co-pathologies,^3^ with small vessel disease being among the most common.^4^ White matter hyperintensities (WMH), visible on T2-weighted MRI, are the most extensively studied imaging markers of small vessel disease.^5^ Although emerging evidence suggests that WMH may result from various pathological processes, they are often assumed to reflect small-vessel-related brain injury associated with underlying vascular risk factors.^6–8^ Regardless of their etiology, multiple studies have shown that the co-occurrence of WMH and AD pathology within an individual accelerates the rate of cognitive decline.^9,10^ Understanding their individual and combined effects on cognitive performance is crucial to determine which cognitive functions are most likely to decline. This understanding, in turn, offers valuable insights into the underlying pathology of the clinical syndrome. However, especially in community-based settings, it remains unclear how effectively WMH burden and AD pathology predict cognitive decline, and which specific cognitive domains are most affected. The recent advancements in blood-based biomarkers for AD pathology provide new opportunities to examine the processes underlying cognitive decline when Aβ and WMHs co-occur, and to evaluate the potential of blood biomarkers as alternatives to imaging-based assessments.

A substantial body of literature has established that vascular cognitive impairment is predominantly characterized by deficits in processing speed and frontal-executive function, whereas AD primarily affects memory.^11^ However, this domain-specific framework represents an oversimplification. Perhaps as a result of their frequent co-occurrence, cognitive decline associated with AD and vascular pathologies frequently affects multiple cognitive domains.^12,13^ For instance, even in preclinical stages, Aβ pathology has been linked to impairments in language and executive function.^14–17^ On the other hand, studies have demonstrated that a higher WMH burden is also linked to faster declines in episodic memory,^18^ some of these findings hold true even after adjusting for hippocampal volume and Aβ-PET burden.^19^ Also blood biomarkers not specific to AD pathology such as neurofilament light chain (NfL) and glial fibrillary acidic protein (GFAP) have been inconsistently linked to declines in a variety of cognitive domains, such as language, executive function, memory, and visuospatial abilities.^20–22^ A thorough comparison of these biomarkers in predicting domain-specific cognitive decline and whether they act additive or synergistic with WMH burden is currently lacking.

In our previous cross-sectional studies, we observed interactions between WMH burden and Aβ on cognitive performance, particularly episodic memory.^8,23^ In this study, we extend those findings by incorporating longitudinal cognitive data and conduct a head-to-head comparison of imaging and blood-based biomarkers in predicting cognitive decline. First, we investigated whether WMH burden predicts domain-specific cognitive decline, and how its predictive value compares with other established AD-related biomarkers, such as Aβ-PET and hippocampal volume, as well as blood-based biomarkers, including plasma p-tau217, GFAP, and NfL. Second, we examined whether cognitive decline associated with these biomarkers is best explained through interactions with WMH burden. Third, given that these biomarkers reflect pathological processes that may lie along a cascade,^24,25^ we aimed to determine which of the identified interactions between biomarkers and WMH best explains cognitive decline. Finally, we considered a scenario in which FLAIR, T1-weighted MRI, and either Aβ-PET or plasma p-tau217 data were available and quantified the proportion of variance in cognitive decline explained by these variables.

## 2. Methods

### 2.1 Study Population

Participants were enrolled in the IDcog study,^26^ a longitudinal, community-based cohort study of cognitive change conducted at the Center for Prevention and Dementia Therapy at the Institute for Regenerative Medicine, University of Zurich, Switzerland. Participants were recruited via newspaper advertisements. To be eligible for enrollment in the study, participants had to be at least 50 years of age and German-speaking. Individuals with comorbid conditions that could interfere with cognitive assessments or MRI and PET procedures were excluded. Individuals with dementia, non-silent strokes, cardiovascular disease resulting in cognitive impairment, clinically significant depression, evidence of cognitive impairment primarily attributable to a medical condition other than potential AD pathology, such as medication use, substance abuse, or heart failure were also excluded.

At baseline, the cohort consisted of 233 participants, all of which underwent Aβ-PET imaging and a cognitive assessment. Participants were followed longitudinally and underwent clinical evaluation and neuropsychological testing at annual assessments. For the current study, we excluded participants without follow up neuropsychological assessment (*n* = 8), with artefacts on the fluid-attenuated inversion recovery (FLAIR) image that did not allow accurate WMH volume quantification (*n* = 7), and with postischemic lesions (*n* = 2).

The study was approved by the local ethics committee (Kantonale Ethikkommission, Zürich, Switzerland) and conducted in accordance with their guidelines and the Declaration of Helsinki. All study participants gave written informed consent.

### 2.2 MRI acquisition and processing

MRI scans were acquired on 3T 750W or Premier scanner models, equipped with either a 32-channel or 48-channel coil scanner (GE Healthcare, Waukesha, WI). A high-resolution 3D T1-weighted fast spoiled gradient recalled (FSPGR) sequence was acquired, with an isotropic voxel size of 0.5 mm, sagittal slice orientation, and repetition time (TR) = 11 ms, echo time (TE) = 5.2 ms, and inversion time (TI) = 600 ms, and a flip angle of 8°. A high-resolution T2-weighted 3D FLAIR was acquired with a voxel size of 0.48*0.48*0.6 mm in a sagittal orientation with imaging parameters of TR/TE/TI = 6502/130/1966 ms, and a flip angle of 90°.

The CAT12 toolbox was used to assess the quality of the T1-weighted images and to extract total intracranial volume (TIV). Following intensity correction and normalization of the T1-weighted images using FreeSurfer, hippocampus volume was segmented using the automatic segmentation of hippocampal subfields (ASHS) software.^27^ All segmentations were visually inspected and manually corrected when necessary. Bilateral hippocampal volume was then calculated as a percentage of TIV.

WMHs were segmented on FLAIR images using the lesion prediction algorithm as implemented in the LST toolbox (www.statistical-modelling.de/lst.html) for SPM as previously described.^28^ Lesion masks were created by binarizing lesion probability maps at a threshold of 0.65. This threshold was selected after applying different thresholds to 20 randomly selected subjects and visually inspecting the generated binarized lesion masks for accuracy. Finally, lesion masks were visually inspected and manually corrected if necessary. We calculated WMH volume as a percentage of TIV. For the path analysis, we divided the cohort into high and low WMH volume groups by stratifying the sample at a WMH volume of 0.25% of the TIV, corresponding to the 66.6th percentile of our cohort.^9^ For the variance decomposition, we used a clinically more meaningful stratification by dividing the cohort based on Fazekas scores for the deep white matter, with a Fazekas score ≥2 indicating higher WMH burden. The threshold of Fazekas score ≥2 was used as it resulted in two groups of adequate and comparable size. Lacunes were found in only six participants and were not considered in any statistical analysis.

### 2.3 Aβ-PET imaging and processing

Dynamic Aβ-PET imaging was performed using a 3T Signa PET/MR scanner (GE HealthCare, Waukesha, WI), with data acquired from 80 to 110 minutes post-injection of approximately 140 MBq [¹⁸F]-flutemetamol.^29^ Image analysis was conducted using PMOD NeuroTool (Version 3.9, PMOD Technologies LLC). A global Aβ standardized uptake value ratio (SUVR) was calculated from the averaged frames acquired between 85-105 minutes post-injection (4 frames of 5 minutes each). The SUVR was derived using a global composite region of interest comprising the bilateral frontal, temporal, and parietal cortices, as well as the anterior and posterior cingulate gyri, normalized to uptake in cerebellar gray matter.^28^ A BRAVO 3D T1-weighted MRI sequence with isotropic voxel size of 1 mm, sagittal slice orientation, TR/TE/TI = 8.4/3.2/450 ms, and flip angle of 12° was acquired in parallel to PET acquisition for gray matter parcellation. A Centiloid threshold of 12 was used to define Aβ positivity, marking the transition from the absence of pathology to the onset of subtle pathological changes.^30^

### 2.4 Plasma biomarker measurement

At the baseline visits, blood was collected from each participant in ethylenediaminetetraacetic acid (EDTA) tubes (Vacutainer EDTA Tubes, BD), inverted 10 times, and centrifuged (1620 × *g*, 12 min, 6°C) to obtain plasma samples. Aliquots of 3 ml plasma were immediately frozen at -80 °C and stored at the Institute for Regenerative Medicine, University of Zurich. Plasma AD biomarker analysis was conducted in the laboratory of Prof. H. Zetterberg and Prof. K. Blennow (Sahlgrenska University Hospital, Mölndal, Sweden). Plasma levels of NfL and GFAP were measured with single-molecule array (SIMOA) technology using the Neurology 4-plex E kit on an HD-X instrument from Quanterix. Plasma Aβ was not considered in the present analysis as its standalone performance in predicting cognitive decline is limited compared to other biomarkers.^2^ Plasma p-tau217 was measured with an in-house Simoa assay developed by the University of Gothenburg. Plasma biomarker levels below the limit of quantitation (LOQ) were set at LOQ/2 according to standard approaches for left-censored LOQ data.^31^ P-tau217 was below the LOQ in 89 participants (41%) and was not available in four participants. Given that chronic kidney disease and higher BMI have been associated with plasma biomarker levels,^32,33^ we included eGFR and BMI as covariates in a sensitivity analysis. eGFR was calculated using the CKD-EPI equation.^34^

### 2.5 Neuropsychological assessment

Cognitive performance was assessed approximately annually. The neuropsychological examination was usually carried out in the morning and lasted around 90 minutes with a short break after 45 minutes. We created five composite scores summarizing the domains of language function, processing speed, executive function, episodic memory, and visuospatial abilities. Language included the animal fluency task (total after 3 minutes) and the 15-word version of the Boston Naming Test. Processing speed included Stroop I (word), Stroop II (color), and TMT part A. Executive function included s-words (correct new words in third minute), digit span backward, Stroop III (color-word), and TMT part B. Episodic memory included immediate recall, delayed recall (words and figures), and recognition from the Consortium to Establish a Registry for Alzheimer’s Disease (CERAD) battery. Visuospatial abilities were assessed using the figure copy tasks in the CERAD test battery (mean score for four figures) and the Rey Complex Figure Task. The mean and standard deviation of cognitively unimpaired individuals who are Aβ negative (Centiloid <12) and have low WMH burden (Fazekas ≤1) were used to calculate z-scores for all time points. Where necessary, we reversed the z-scores of tests so that a higher score represents better performance across all tests. A global cognitive summary score was also created by averaging all individual cognitive tests, including the MMSE.

### 2.6 Statistical Analysis

We investigated three imaging-based (Aβ SUVR, hippocampus volume, WMH volume) and three blood-based (p-tau217, NfL, GFAP) biomarkers. We first used partial Pearsons’s correlation analyses to investigate the correlations among the biomarkers. Correlations were first adjusted for sex and then for sex and age. All biomarkers were log-transformed for this analysis, except hippocampal volume, which followed a normal distribution.

We ran a series of linear mixed-effect models with subject-specific random intercept and random slopes to examine the longitudinal associations between these six biomarkers and decline in cognitive domains as well as the global composite score. Three sets of linear mixed-effects models were examined. First, we ran separate models for each biomarker. Estimates for biomarker associations with cognition at baseline were also derived from these separate linear models. Second, we ran combined models that included all biomarkers simultaneously, allowing us to assess the independence of each predictor. Variance inflation factors (VIFs) for all biomarkers were ≤2.36, indicating no major concerns with multicollinearity. Third, we examined an interaction model that included the three-way interaction between each biomarker, WMH volume, and time. Likelihood ratio tests were used to determine whether including the three-way interaction improved the model fit compared to a simpler model that includes only the interactions with time. *P* values of the likelihood ratio tests were corrected using false discovery rate (FDR) correction. All models were adjusted for baseline age, sex, years of education, APOE ε4 status, and their interactions with time. To enable direct comparison of estimates, all biomarkers were z-transformed. For simplicity and ease of result visualization and comparison, we assumed a linear decline across all cognitive domains. The trajectories of the Clinical Dementia Rating Scale Sum of Boxes (CDR-SOB) were analyzed using linear mixed-effects models, with time modeled as natural cubic splines with two degrees of freedom.^35^ Due to the small number of individuals who progressed in the CDR-SOB, participants were divided into four groups for this analysis based on Aβ-PET and WMH status: Aβ- WMH-, Aβ- WMH+, Aβ+ WMH-, and Aβ+ WMH+.

Given the identified interactions between biomarkers and WMH on cognitive decline, and the correlations among biomarkers, we next sought to determine which interaction was decisive. To do this, we used multigroup path analysis and separated the sample into high and low WMH groups (stratified at the 66.6th percentile). This approach had the advantage that all interactions between WMH groups and biomarkers could be modeled simultaneously. We specifically focused on episodic memory performance as interactions between biomarkers and WMH burden were particularly pronounced for this cognitive domain and were observed for all biomarkers except NfL. Individual slopes of episodic memory change were estimated and extracted from linear mixed-effects models with random intercepts and slopes per participants and time as the sole predictor. Based on literature suggesting that Aβ, GFAP, p-tau217, and hippocampus volume become abnormal in a sequential manner,^24,25^ we specified our model accordingly. NfL was not included in this model because it did not interact with WMH to predict episodic memory decline. As only paths predicting episodic memory change were of interest in the present analysis, only these paths were allowed to vary by WMH group whereas paths among biomarkers were estimated using the combined sample. Each path was then sequentially constrained, and model fit was compared to the unconstrained model using a likelihood ratio test. Significant differences indicated that paths differ between groups and should be estimated separately. A final multigroup model was estimated with 1000 bootstrap samples.

We estimated the percentage of variance (R²) in episodic memory slopes that can be attributed to the biomarkers Aβ SUVR, plasma p-tau217, and hippocampus volume using commonality analysis. By fitting regression models across all possible combinations of predictors, this method divides the total R² into components that are uniquely attributable to each predictor, as well as variance that is shared among predictors. Aβ SUVR was selected due to its strong association with cognitive decline, while hippocampal volume was included because of its independent contribution to cognitive decline. Plasma p-tau217 was chosen as it is currently considered the most promising fluid-based biomarker.^2^ We examined three models: one including Aβ SUVR and hippocampal volume, one including plasma p-tau217 and hippocampal volume, and one including all three biomarkers. All models controlled for age, sex, and education. Analyses were conducted in the total cohort as well as separately for participants with Fazekas scores ≥2 and <2.

Statistical significance was set at *P* < 0.05 (two-sided). Path analysis results were considered significant when the 95% bootstrap confidence interval did not include zero. Analyses were performed in R v4.4.1 using the packages *nlme* (v3.1.165) and *effects* (v4.2.2) for linear mixed-effect models, *lavaan* (v0.6.18) for the path analysis, and *rsq* (v2.7) package for the variance decomposition.

## 3. Results

### 3.1 Cohort characteristics

Table 1 summarizes the characteristics of the study cohort. A total of 216 participants were included in the present analysis. All participants had baseline Aβ-PET imaging and WMH volume data, as well as longitudinal cognitive assessments. The mean age at baseline was 66.9 years; 103 participants (47.7%) were female, 51 (23.6%) were carriers of an APOE ε4 allele, and 67 (31.0%) had a Centiloid value greater than 12 and were classified as Aβ positive. The median follow-up duration was 5.3 years, with the number of cognitive assessments per participant ranging from two to eight.

**Table 1.** Baseline cohort characteristics.

|  | **Total (N = 216)** |
| --- | --- |
| Age at Baseline NP, median years [range] | 67.2 [51-90] |
| Education, mean years (SD) | 15.4 (2.8) |
| Female sex, n (%) | 103 (47.7) |
| MMSE, mean (SD) | 29.2 (1.1) |
| APOE ε4 carriers, N (%) | 51 (23.6) |
| CDR global of 0.5, N (%) | 27 (12.7) |
| CDR Sum of Boxes ≥0.5, N (%) | 28 (13.0) |
| Aβ-PET burden, SUVR, median (IQR) | 1.21 [1.15-1.29] |
| Centiloid >12, N (%) | 67 (31.0) |
| Time difference Baseline NP and Aβ-PET, days, median (IQR) | 18.5 [9-35] |
| Plasma p-tau217<sup>a</sup>, pg/ml, median (IQR) | 1.88 [1.44-2.37] |
| Plasma GFAP, pg/ml, median (IQR) | 109.1 [73.2-130.3] |
| Plasma NfL, median (IQR) | 19 [14-25.1] |
| Hippocampus Volume, % of TIV, mean (SD) | 0.45 (0.04) |
| WMH Volume, % of TIV, median (IQR) | 0.16 [0.08-0.35] |
| Fazekas score, 0/1/2/3, N (%) | 31/98/56/25 (14.8/46.7/26.7/11.9) |
| Number of NP assessments, 2/3/4/5/>6, N | 10/26/39/71/70 |
| Follow-up years, median years (IQR) | 5.26 [5.0-6.75] |

<sup>a</sup>Values below the limit of detection excluded.
Abbreviations: IQR = interquartile range, NP = neuropsychological, PET = positron emission tomography, SD = standard deviation, SUVR = standardized uptake value ratio.

### 3.2 Biomarker correlations

Figure 1 plots pair-wise Pearson’s correlations among the investigated biomarkers. Significant correlations were observed among all biomarkers when adjusting only for sex (Figure 1A). After additional adjustment for age, the strength of these correlations was markedly reduced, and only the associations among Aβ SUVR, p-tau217, NfL, and GFAP remained significant (Figure 1B).

**Figure 1.**
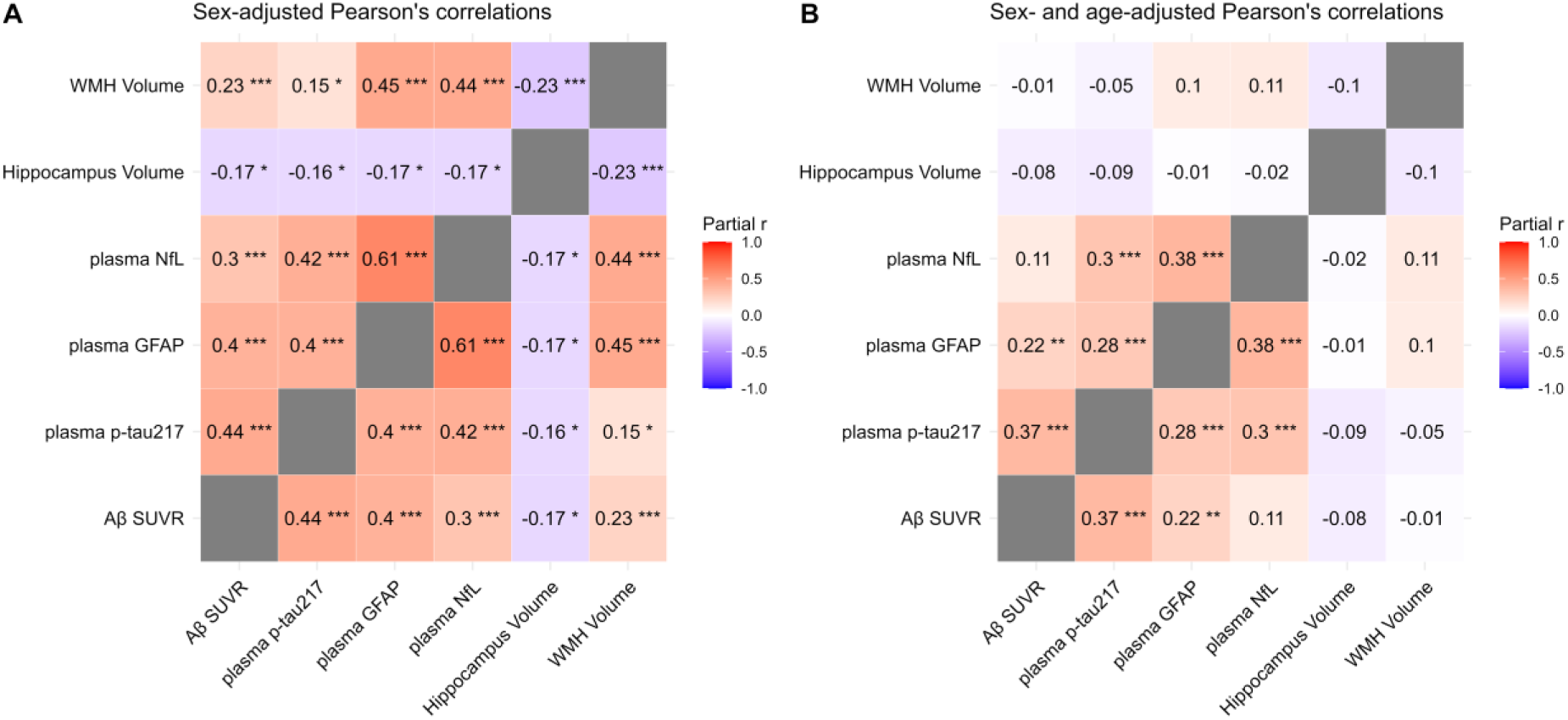
Partial Pearson’s correlations among the investigated biomarkers. All biomarkers, except hippocampus volume, were log-transformed for this analysis. \**P* < 0.05, \*\**P* < 0.01, \*\*\**P* < 0.001.

### 3.3 WMH volume moderates biomarker-related cognitive decline

Using separate linear mixed-effects models for each biomarker, we found that all investigated biomarkers predicted cognitive decline to varying degrees (Figure 2A). Each biomarker was associated with a decline in the global cognitive composite score, with Aβ SUVR showing the largest effect size. Aβ SUVR, plasma p-tau217, and plasma GFAP emerged as the strongest predictors of decline in both episodic memory and executive functions (all *P* < 0.001), with larger effect sizes observed for episodic memory. Higher plasma NfL levels were also associated with decline in both domains (episodic memory: p = 0.011; executive function: *P* = 0.043), whereas a smaller hippocampus volume predicted faster decline in episodic memory (*P* = 0.016) but showed only a non-significant trend in executive functions (*P* = 0.051). WMH volume predicted decline in executive functions (*P* = 0.005) but not in episodic memory (*P* = 0.57). Effect sizes were generally smaller for the language, processing speed, and visuospatial domains. Higher Aβ SUVR (*P* = 0.022), and marginally p-tau217 (*P* = 0.061) and hippocampal volume (*P* = 0.062), were associated with faster language decline. Except for Aβ SUVR (*P* = 0.24), all biomarkers predicted decline in processing speed (hippocampus volume: *P* = 0.001; p-tau217: P = 0.01; WMH volume: *P* = 0.022; GFAP: *P* = 0.028; NfL: *P* = 0.036). No biomarkers significantly predicted decline in visuospatial ability, with plasma p-tau217 being closest to the significance threshold (*P* = 0.055). At baseline (i.e., cross-sectionally), only few significant associations between biomarkers and cognitive performance were observed (Figure S1): Aβ SUVR was associated with episodic memory (β = -0.16, *P* = 0.035); GFAP was associated with global cognition (β = -0.15, p = 0.003), episodic memory (β = -0.15, p = 0.05), processing speed (β = -0.15, *P* = 0.023), and language (β = -0.18, *P* = 0.016); NfL was associated with global cognition (β = -0.10, p = 0.046); and hippocampus volume was associated with episodic memory (β = 0.16, *P* = 0.01).

**Figure 2.**
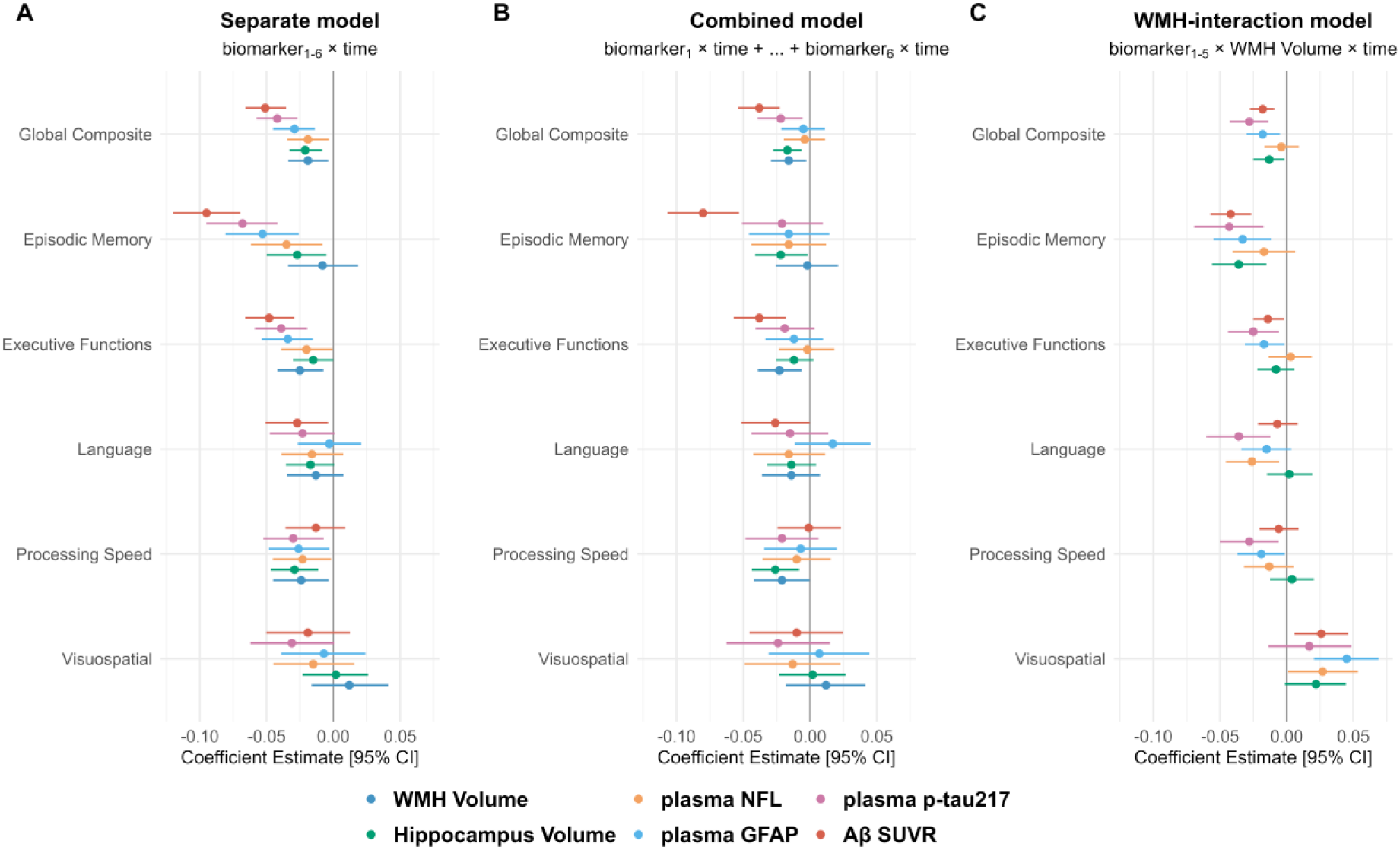
Head-to-head comparison of imaging and plasma biomarkers in predicting global and domain-specific cognitive decline. **(A)** shows estimates from separate linear mixed-effects models for each biomarker. **(B)** shows estimates from a combined model including all biomarkers simultaneously. **(C)** shows estimates from separate linear mixed-effects models for the interaction effects between each biomarker and WMH volume. Error bars represent 95% confidence intervals. All variables were standardized and estimates for hippocampus volume were reversed to allow direct comparison of effect sizes. All models were adjusted for age, sex, APOE ε4 status, education, and their interactions with time.

When all biomarkers were included in the same linear mixed-effects model (Figure 2B), Aβ SUVR emerged as the strongest predictor of cognitive decline in the global composite (*P* < 0.001), episodic memory (*P* < 0.001), executive function (*P* < 0.001), and language (*P* = 0.048) domains. Plasma p-tau217 (*P* = 0.009), hippocampal volume (*P* = 0.002), and WMH volume (p = 0.018) also remained significantly associated with decline in the global composite score, suggesting they provide independent predictive value. Next to Aβ SUVR, hippocampus volume was additionally identified as an independent predictor of decline in episodic memory (*P* = 0.033), whereas WMH volume was identified as an independent predictor of executive function decline (*P* = 0.007). For the processing speed domain, hippocampus volume (*P* = 0.004) and WMH volume (*P* = 0.046) both significantly predicted decline. No biomarker showed a significant association with decline in visuospatial abilities in this combined model.

For each cognitive domain, we identified significant interactions between at least two biomarkers and WMH volume in predicting cognitive decline (Figure 2C). Interactions were particularly pronounced for episodic memory decline. These interactions consistently showed that elevated levels of both WMH volume and a given biomarker synergistically predicted faster decline. The exception was the visuospatial abilities domain, for which interactions indicated that higher levels of Aβ SUVR, GFAP, and NfL predict visuospatial decline when WMH volume is low, but less so when WMH volume is high (Figure S2). Notably, all models incorporating significant three-way interactions (i.e., time × biomarker × WMH volume) demonstrated a significantly better fit compared to simpler models that included only the two-way interactions with time (i.e., time × biomarker + time × WMH volume) (Table 2). This was supported by, in some cases substantial, reductions in AIC. The overall increase in R² when the three-way interaction term was included was modest for most cognitive domains. The greatest improvement in predicting change in cognitive performance was observed for episodic memory, with the interaction model of Aβ SUVR accounting for an additional 14.3% of the variance.

**Table 2.** Model fit comparison between two-way (time × biomarker + time × WMH volume) and three-way (time × biomarker × WMH volume) interaction models.

| Outcome | Biomarker | ΔAIC | ΔR<sup>2</sup> | R<sup>2</sup> (three-way interaction model) | FDR p value |
| --- | --- | --- | --- | --- | --- |
| Episodic Memory | Aβ SUVR | -74.55 | 0.1431 | 0.477 | <0.001 |
| Global Composite | Aβ SUVR | -29.75 | 0.0656 | 0.4679 | <0.001 |
| Episodic Memory | Hippocampus Volume | -19.37 | 0.0513 | 0.3435 | <0.001 |
| Episodic Memory | plasma p-tau217 | -25.09 | 0.0429 | 0.3359 | <0.001 |
| Episodic Memory | plasma GFAP | -18.1 | 0.0398 | 0.3286 | <0.001 |
| Global Composite | plasma GFAP | -17.54 | 0.0314 | 0.4349 | <0.001 |
| Visuospatial | plasma GFAP | -21.07 | 0.0284 | 0.133 | <0.001 |
| Global Composite | Hippocampus Volume | -7.54 | 0.0277 | 0.417 | 0.0103 |
| Global Composite | plasma p-tau217 | -20.13 | 0.0201 | 0.4203 | <0.001 |
| Executive Functions | Aβ SUVR | -5.72 | 0.015 | 0.3341 | 0.0195 |
| Processing Speed | plasma GFAP | -4.71 | 0.014 | 0.2995 | 0.0256 |
| Executive Functions | plasma GFAP | -5.79 | 0.0105 | 0.3289 | 0.0195 |
| Visuospatial | Aβ SUVR | -5.02 | 0.0091 | 0.107 | 0.0236 |
| Visuospatial | plasma NFL | -3.42 | 0.008 | 0.1069 | 0.0385 |
| Processing Speed | plasma p-tau217 | -5.1 | 0.0065 | 0.2864 | 0.0236 |
| Language | plasma p-tau217 | -7 | 0.0045 | 0.2288 | 0.0123 |
| Language | plasma NFL | -4.34 | 0.0042 | 0.2337 | 0.0272 |
| Executive Functions | plasma p-tau217 | -4.54 | 0.0004 | 0.3183 | 0.0262 |

Shown are only parameters for models where there was a significant three-way interaction, but FDR correction was conducted across p values from all model comparisons.

Detailed estimates for all models shown in Figure 2 can be found in Tables S1-S3. Results were unchanged when we additionally controlled for BMI and eGFR (Figure 3) or hypertension (Figure 4). No interaction between Aβ SUVR, hypertension, and time on episodic memory decline was observed (β = 0.002, *P* = 0.94). The results were attenuated but remained similar when participants with a baseline CDR global >0 were excluded (Figure 5). However, interactions between biomarkers and WMH volume were primarily observed for Aβ SUVR.

**Figure 3.**
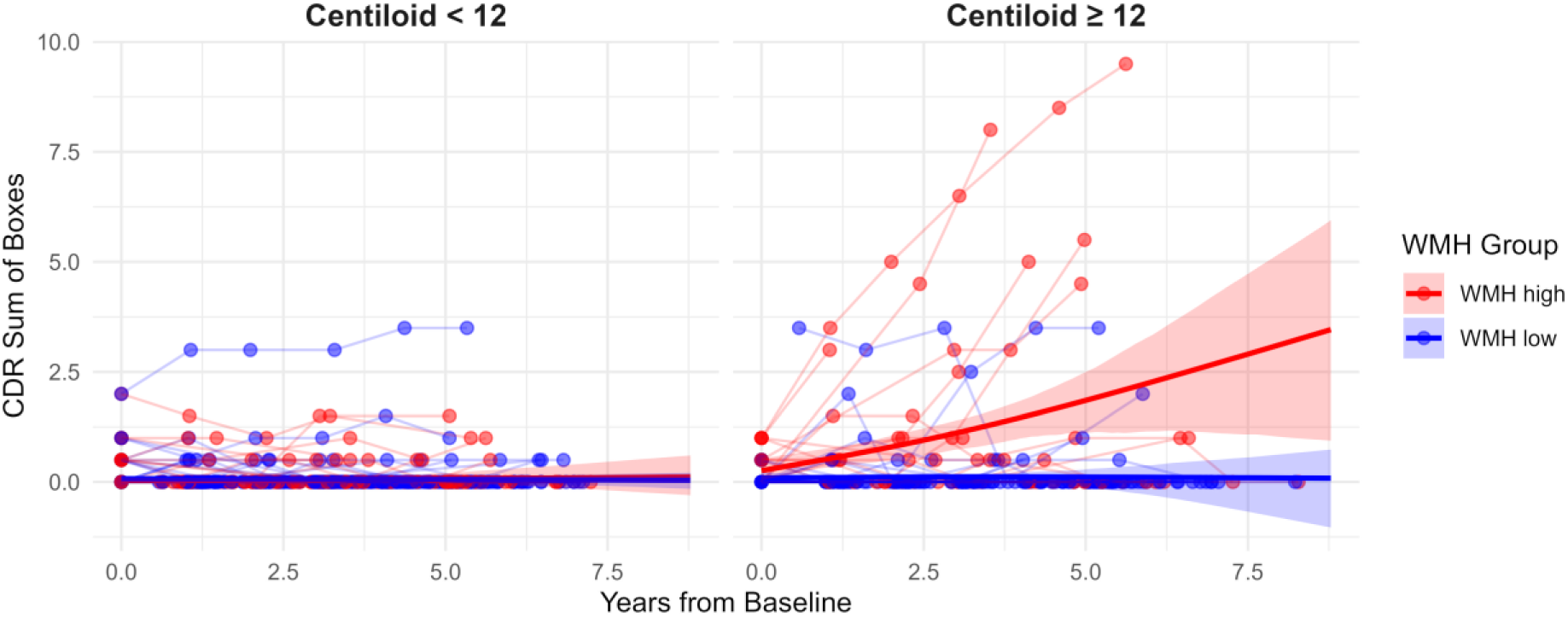
Progression in the CDR-SOB is only seen in individuals with both increased Aβ-PET burden and increased WMH burden. Shaded areas represent 95% confidence intervals, derived from 1000 bootstrap samples.

**Figure 4.**
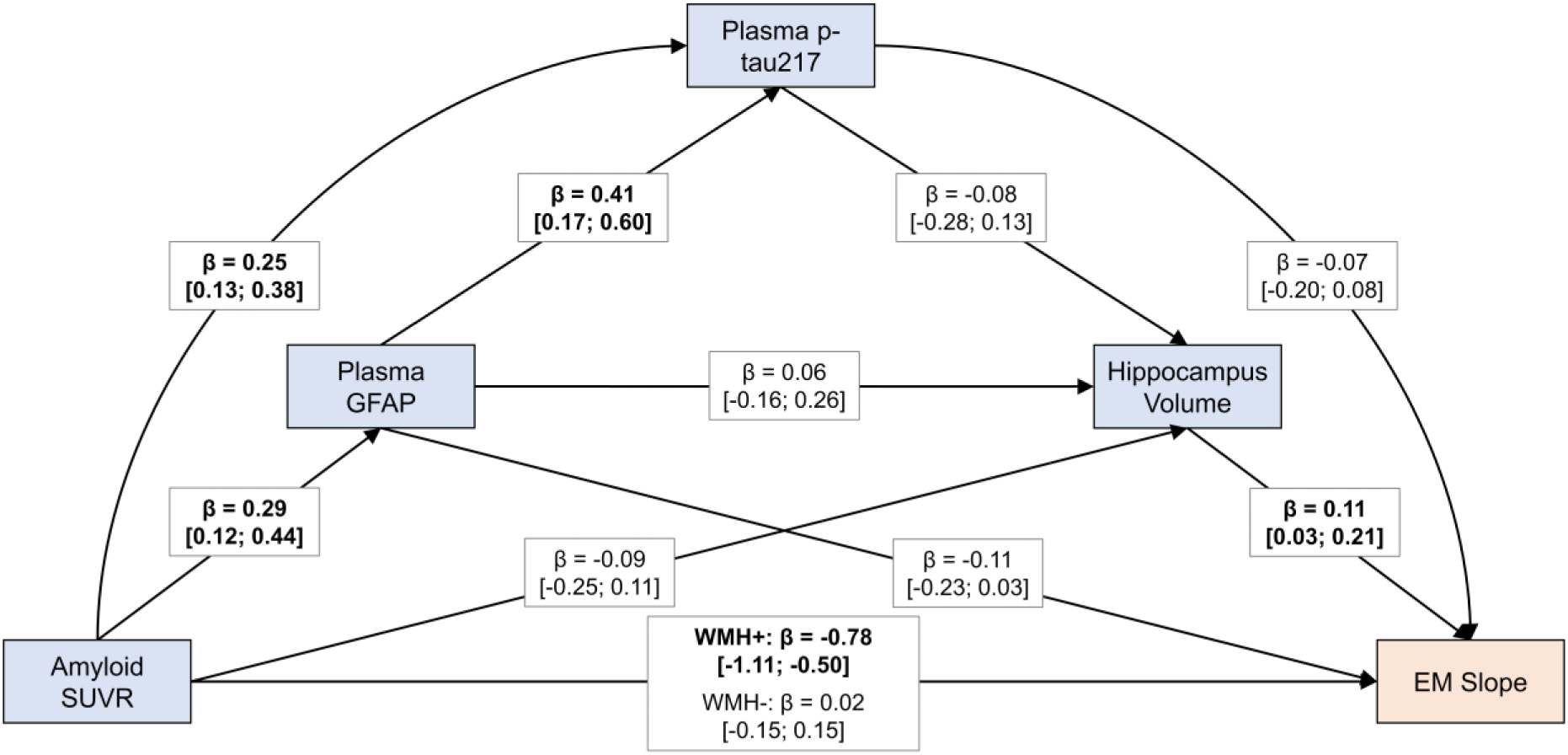
The interaction between amyloid PET and WMH burden is the primary interaction associated with faster episodic memory decline. Shown is the final multigroup path model allowing only paths to vary by WMH group for which significant group differences were observed. All paths were adjusted for age, sex, and APOE ε4 status. Paths to EM (episodic memory) slope were additionally adjusted for education. All variables were standardized prior to model entry. Values in brackets indicate 95% confidence intervals, derived from 1000 bootstrap samples. Bolded values indicate statistically significant associations (confidence interval excludes 0).

**Figure 5.**
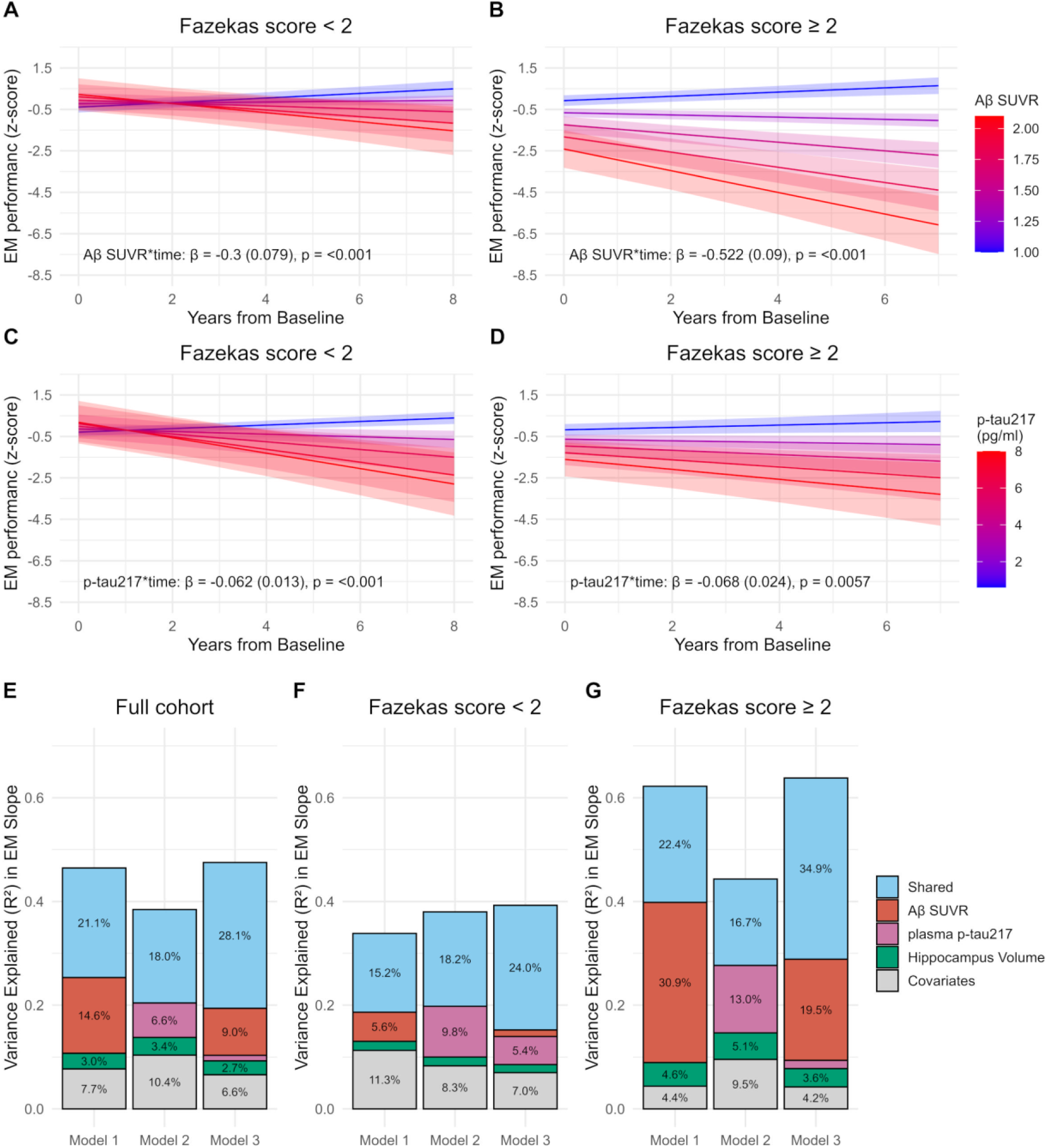
Prediction of episodic memory slope in groups with low and high Fazekas score. **A-D** show the predicted EM (episodic memory) z-score over time for a given baseline level of Aβ SUVR **(A and B)** or plasma p-tau217 **(C and D)**. Estimates are from separate linear mixed-effects models in groups with low and high Fazekas scores. E-G illustrate the results of the variance decomposition for the full cohort **(E)** as well as for individuals with low **(F)** and high **(G)** Fazekas scores. Shared indicates the proportion of variance that was shared with one or more variable in the model. For Aβ SUVR, plasma p-tau217, and hippocampus volume the variance that is unique to these predictors in each model is shown. For covariates, the variance unique to age, sex, and education was combined.

In line with the results from the cognitive domains, decline as measured using the CDR-SOB was observed only in the group of participants with both elevated Aβ-PET burden and high WMH volume (Figure 3).

### 3.4 Multigroup path analysis

In the analyses above, we demonstrated that individuals with elevated WMH volume showed significantly faster cognitive decline for a given level of Aβ SUVR, plasma p-tau217, plasma GFAP, and hippocampal volume compared to individuals with lower WMH volume. These interaction effects were strongest for episodic memory. Given the correlations among these biomarkers (see Figure 1), we next used multigroup path analysis to determine which interaction primarily drives cognitive decline.

Constraining the path from Aβ SUVR to episodic memory slope significantly worsened model fit (*P* < 0.0001, ΔAIC = 33.3), suggesting that this path differs between high and low WMH groups. In contrast, constraining the paths from GFAP (*P* = 0.16, ΔAIC = 0), p-tau217 (*P* = 0.15, ΔAIC = 0.1), or hippocampal volume (*P* = 0.09, ΔAIC = 0.9) to episodic memory slope did not significantly decrease the model fit. Therefore, the final multigroup path model was estimated with the path from Aβ SUVR to episodic memory slope varying by WMH group and all remaining paths were estimated using the combined sample. This model is shown in Figure 4.

As expected, the model revealed significant correlations among Aβ SUVR, plasma GFAP, and plasma p-tau217. None of these biomarkers were significantly associated with hippocampal volume. Regarding episodic memory slope, a strong association was found between higher Aβ SUVR and steeper episodic memory decline in individuals with high WMH burden, but not in those with low WMH burden. GFAP and p-tau217 were not significantly associated with episodic memory slope after accounting for the Aβ SUVR × WMH group interaction. Hippocampal volume emerged as an independent predictor of episodic memory decline, regardless of the WMH group.

Because interactions between all biomarkers (except NfL) and WMH burden were also observed for global cognition, we repeated the analysis using global cognition slope as the outcome. Results mirrored those from the episodic memory analysis (Figure S6). Although significant WMH group differences were found for the paths from both Aβ SUVR and plasma GFAP to global cognition, only the Aβ SUVR path in the high WMH group was significant in the final multigroup path model (WMH+: β = -0.46, 95% CI: -0.86 to -0.16; WMH-: β = -0.05, 95% CI: -0.24 to 0.09). Additionally, lower hippocampal volume (β = 0.12, 95% CI: 0.02 to 0.23) and higher p-tau217 (β = -0.16, 95% CI: -0.32 to -0.01) levels were independently associated with faster global cognitive decline regardless of the WMH group.

### 3.5 Variance decomposition in samples stratified by Fazekas score

The visual Fazekas rating scale aligned well with quantitative WMH volumes: all pairwise comparisons between Fazekas score groups were statistically significant (Wilcoxon rank-sum tests, *P* < 0.05), indicating a consistent stepwise increase in WMH burden across severity levels (Figure S7). In a linear-mixed effects model, Fazekas score interacted with Aβ SUVR and showed a stepwise decrease in cognitive performance with increasing scores (Table S4).

We then stratified the cohort by Fazekas score (cut-off ≥2) and tested in separate samples the performance of Aβ SUVR, plasma p-tau217, and hippocampus volume to predict episodic memory decline. As illustrated in Figure 5A and 5B, Aβ SUVR predicts decline in both samples, but the decline is much more pronounced in the groups with Fazekas ≥2 compared to the group with Fazekas <2. A similar observation was made for plasma p-tau217 (Figure 5C and 5D), although compared to Aβ-PET, plasma p-tau217 appeared to be a stronger predictor of episodic memory decline in the group with Fazekas <2. Variance decomposition reveals that, in the full cohort, Aβ SUVR contributes more than twice the unique variance to the episodic memory slope compared to plasma p-tau217 (Figure 5E). When both biomarkers are included in the same model, p-tau217 adds minimal unique variance. In the low Fazekas score group, p-tau217 explains more unique variance than Aβ SUVR (Figure 5F). Conversely, in the high Fazekas group, Aβ SUVR accounts for substantial unique variance, while the contribution from plasma p-tau217 is comparatively smaller (Figure 5G). When both markers are included together, the unique variance explained by p-tau217 is negligible, suggesting the predictive value of p-tau217 is captured fully in the Aβ SUVR variable.

## 4. Discussion

In this study, we examined the combined and independent contributions of WMHs and AD-related biomarkers, including both imaging- and blood-based markers, in predicting longitudinal cognitive decline in a well-characterized, community-based cohort. Our findings reveal that WMHs independently predict decline in executive functions, processing speed, and global cognition, but also moderate the effects of other biomarkers on almost all investigated domains. Individual biomarkers including Aβ-PET, hippocampal volume, plasma p-tau217, GFAP, and NfL are significant predictors of decline across multiple cognitive domains and predict cognitive decline when WMH burden is low, but the biomarker-related decline is markedly increased when WMH burden is high. This interaction was particularly pronounced for episodic memory decline, highlighting the importance of cerebrovascular pathology in modulating AD-related cognitive consequences. Our results provide further evidence that vascular pathological burden is a key player in driving cognitive decline in preclinical and early AD and contributes to the heterogeneity observed in cognitive trajectories. Therefore, considering Aβ-PET and blood-based biomarkers in the context of an individual’s WMH burden may improve the prognostic value of these biomarkers.

These results strongly suggest that the co-occurrence of WMHs and AD pathology accelerates cognitive decline beyond the effects of either pathology alone. The synergistic effect was particularly evident in changes in the CDR-SOB, where only individuals with elevated levels of both pathologies showed noticeable progression. Therefore, coexisting cerebrovascular disease may shorten the preclinical stage of AD and preventing or mitigating vascular pathology may delay disease progression.

The extent to which interactions between AD biomarkers and WMH lesions can be detected may depend on the stage of the disease and may be more pronounced in the early stages. It has been reported that the exacerbating effects of vascular brain injury on the relationship between AD pathology and cognition is pronounced in early disease stages, when tau pathology is still confined to the medial temporal lobe regions, but when tau pathology extends into limbic or neocortical regions, it appears to have little additional impact on cognitive deficits.^36^ This is consistent with other studies indicating that AD pathology becomes the primary driver of dementia risk in later disease stages.^37–39^ Our results would support this disease stage-specific effects of AD and vascular pathologies. Similarly, Ferrari-Souza found that in a cognitively unimpaired sample from the Alzheimer’s Disease Neuroimaging Initiative (ADNI), AD pathology interacts with WMH burden to accelerate longitudinal cognitive decline.^40^ Nevertheless, even in cognitively unimpaired samples, longitudinal studies do not always support an interactive association between AD pathology and vascular brain pathologies on cognitive decline.^9,19^

In models including all biomarkers, Aβ-PET, p-tau217, WMH burden, and hippocampal volume independently predicted decline in the global composite score, while domain-specific cognitive decline was exclusively associated with imaging biomarkers, indicating that imaging biomarkers are still superior to plasma biomarkers in detecting subtle cognitive decline. These combined models also clearly delineated characteristic pathology-cognition profiles: Aβ-PET was strongly associated with decline in episodic memory, and to a lesser extent with executive function and language; WMH burden was related to decline in executive functions and processing speed. Given its well-documented role in memory function,^41^ it is not surprising that hippocampus volume provided unique information for the prediction of episodic memory change. However, it was surprising to discover that the hippocampus emerged as the strongest predictor of processing speed, even though cross-sectional studies reported such an association before.^42,43^ Notably, this association persisted even when the analysis was limited to individuals with a CDR of 0 (see Figure S5), suggesting that the hippocampus plays a central role in cognitive aging, not only in episodic memory, but also in other cognitive domains.^44^

Among the plasma biomarkers evaluated, p-tau217 emerged as the most promising predictor of cognitive decline. Plasma p-tau217 showed domain-specific associations similar to those of Aβ-PET, supporting its role as a promising biomarker relatively specific for AD pathology.^2^ Plasma p-tau217 was also the only biomarker that marginally predicted change in visuospatial abilities. Consistent with prior research,^20,22^ plasma NfL and GFAP predicted decline across multiple domains. However, their ability to predict cognitive decline was limited. Both biomarkers showed the weakest effect sizes in separate models and did not predict decline in any of the combined models. In addition, NfL showed little to no interaction with WMH volume. Collectively, these findings suggest that plasma NfL and GFAP may have limited utility for predicting cognitive decline in community-based cohorts and imaging biomarkers or plasma p-tau217 may be preferred when it comes to predicting the rate of cognitive decline.

Model fit was consistently improved across cognitive domains by taking into account the interaction between biomarkers and WMH burden. Although interactions were strongest for episodic memory, the lack of domain specificity is not entirely surprising given the interdependence of cognitive processes. Salthouse proposed that decreases in processing speed have a fundamental role in cognitive aging that causes broader declines in other cognitive domains as many higher-order tasks depend on basic processing speed.^45^ Conversely, while WMH burden amplifies AD-related cognitive decline, AD pathology, particularly p-tau217, also appeared to exacerbate more vascular pathology-driven processing speed deficits, highlighting the bidirectional nature of these interactions. On a more pathophysiological level, white matter injury to specific white matter tracts plays a critical role in memory differences seen in older adults^46,47^ and may exacerbate the memory decline associated with tau pathology in the medial temporal lobe. In our previous cross-sectional work, we found that the interaction between Aβ and WMH on episodic memory performance were especially pronounced when lesions were located in periventricular regions,^8^ which contains long-range fibers that are essential for many cognitive processes. Furthermore, WMH burden may partially reflect the extent of cerebral amyloid angiopathy (CAA),^48,49^ which is frequently linked to WMH burden in the posterior brain.^50,51^ Notably, it has been found that the interaction between CAA and Aβ plaques predicts higher tau pathology at postmortem^52^ and faster tau accumulation on PET imaging in cognitively unimpaired individuals.^53^ These findings suggest that several mechanisms may be at play: lesions around the ventricles may affect long-range white matter connections and thereby amplify cognitive consequences of functional and structural deterioration of AD pathologies,^54^ and lesions in posterior brain regions may be related to the degree of CAA which in interaction with Aβ plaques causes faster tau accumulation thereby promoting faster cognitive decline.^52^ Longitudinal studies involving regional WMH together with AD pathology are crucial to better understand when and why these pathologies interact to accelerate cognitive decline.

Our multigroup path analysis showed that although all biomarkers except plasma NfL interacted with WMH burden to predict faster decline in episodic memory and global cognition, the interaction between Aβ-PET and WMH was the primary predictor of decline. Once this interaction was accounted for, no other biomarker-WMH interaction was significant. The model also revealed that hippocampal volume was an independent predictor of memory and global cognitive decline, regardless of WMH burden, suggesting that white matter lesions may amplify the impact of AD pathology before neurodegenerative changes occur. Importantly, accounting for Aβ SUVR × WMH interaction explained an additional 14% variance in episodic memory change in our linear-mixed effects model. The explained variance also increased markedly in our variance decomposition. Applying a simple dichotomization at Fazekas score 2, Aβ SUVR accounted for approximately 25% more unique variance in the episodic memory slope among individuals with Fazekas ≥2 compared to those with Fazekas <2. Plasma p-tau217 displayed a similar but notably weaker trend, contributing only about 3.5% more unique variance among individuals with Fazekas ≥2 compared to those with Fazekas <2. This suggests the interaction may be more specific to Aβ pathology than to soluble p-tau, but it is unclear if this is due to variations in the underlying pathological mechanisms or measurement specificity.

A limitation of this study is that our cohort may be not representative of the general population. Participants were exclusively White and recruited through newspaper ads, suggesting they might be more health-conscious and motivated. Therefore, future studies that include cohorts with more diverse ethnic, racial, and socioeconomic backgrounds are warranted. Although we imputed missing values below the limit of quantitation, plasma p-tau217 levels were below the limit of detection in 89 participants (41%), which may have affected the accuracy of our estimates. Finally, cognitive symptoms due to vascular pathologies may vary by lesion number, type, location, and combinations of vascular pathologies. However, as we excluded patients with infarcts and only six participants had lacunes, our sample is relatively homogeneous with regards to vascular brain lesions, and the observed moderating effects likely reflect WMH burden specifically.

In conclusion, our results provide strong support for an interactive association of Aβ and WMHs on cognitive decline, particularly episodic memory but also in other cognitive domains. Aβ-PET was the main factor interacting with WMHs to predict episodic memory decline. However, in models excluding Aβ-PET, significant interactions were also observed for hippocampal volume, plasma p-tau217, and GFAP, though to a lesser extent for NfL. Considering Aβ-PET or blood-based biomarkers together with WMH burden may improve the prognostic accuracy for disease progression.

## Data Availability

All data produced in the present study are available upon reasonable request to the authors.

## 6. Acknowledgements

The authors would like to thank the entire IDcog study team, including the staff of the PET/CT/MR Center at the Department of Nuclear Medicine, University Hospital Zurich and the database team of the Institute for Regenerative Medicine, University of Zurich. Special gratitude to the IDcog participants and their study partners, without whom this study would not have been possible.

## 9. Consent statement

All participants provided written informed consent.

