## Supplementary material for "White Matter Hyperintensities Impact Biomarker Prediction of Cognitive Decline": Table S1

**Supplementary Table 1 Results of the separate linear mixed effects models.**

| Outcome | Predictor | Estimate | CI Lower | CI Upper | *P* value | Marginal R2 | Conditional R2 |
| --- | --- | --- | --- | --- | --- | --- | --- |
| Global Composite | Aβ SUVR | -0.051 | -0.066 | -0.035 | <0.0001 | 0.38 | 0.92 |
| Executive Functions | Aβ SUVR | -0.048 | -0.066 | -0.029 | <0.0001 | 0.28 | 0.86 |
| Episodic Memory | Aβ SUVR | -0.095 | -0.12 | -0.07 | <0.0001 | 0.32 | 0.82 |
| Processing Speed | Aβ SUVR | -0.013 | -0.036 | 0.009 | 0.2425 | 0.26 | 0.85 |
| Language | Aβ SUVR | -0.027 | -0.051 | -0.004 | 0.0222 | 0.21 | 0.77 |
| Visuospatial | Aβ SUVR | -0.019 | -0.05 | 0.013 | 0.2394 | 0.1 | 0.42 |
| Global Composite | plasma p-tau217 | -0.042 | -0.057 | -0.027 | <0.0001 | 0.38 | 0.92 |
| Executive Functions | plasma p-tau217 | -0.039 | -0.059 | -0.019 | 0.0001 | 0.28 | 0.86 |
| Episodic Memory | plasma p-tau217 | -0.068 | -0.095 | -0.042 | <0.0001 | 0.28 | 0.82 |
| Processing Speed | plasma p-tau217 | -0.03 | -0.052 | -0.007 | 0.01 | 0.27 | 0.85 |
| Language | plasma p-tau217 | -0.023 | -0.048 | 0.001 | 0.061 | 0.22 | 0.77 |
| Visuospatial | plasma p-tau217 | -0.031 | -0.062 | 0.001 | 0.0551 | 0.1 | 0.43 |
| Global Composite | plasma GFAP | -0.029 | -0.045 | -0.014 | 0.0003 | 0.39 | 0.92 |
| Executive Functions | plasma GFAP | -0.034 | -0.053 | -0.015 | 0.0004 | 0.29 | 0.86 |
| Episodic Memory | plasma GFAP | -0.053 | -0.081 | -0.026 | 0.0002 | 0.28 | 0.82 |
| PS/Attention | plasma GFAP | -0.026 | -0.048 | -0.003 | 0.0278 | 0.28 | 0.85 |
| Language | plasma GFAP | -0.003 | -0.026 | 0.021 | 0.8194 | 0.22 | 0.77 |
| Visuospatial | plasma GFAP | -0.007 | -0.039 | 0.024 | 0.6449 | 0.1 | 0.43 |
| Global Composite | plasma NFL | -0.019 | -0.034 | -0.003 | 0.0181 | 0.36 | 0.92 |
| Executive Functions | plasma NFL | -0.02 | -0.039 | -0.001 | 0.0425 | 0.28 | 0.86 |
| Episodic Memory | plasma NFL | -0.035 | -0.062 | -0.008 | 0.0111 | 0.25 | 0.82 |
| Processing Speed | plasma NFL | -0.023 | -0.045 | -0.002 | 0.0356 | 0.27 | 0.85 |
| Language | plasma NFL | -0.016 | -0.039 | 0.008 | 0.1867 | 0.22 | 0.77 |
| Visuospatial | plasma NFL | -0.015 | -0.045 | 0.016 | 0.3485 | 0.1 | 0.43 |
| Global Composite | Hippocampus Volume | 0.021 | 0.008 | 0.033 | 0.0011 | 0.37 | 0.92 |
| Executive Functions | Hippocampus Volume | 0.015 | 0 | 0.03 | 0.0509 | 0.27 | 0.86 |
| Episodic Memory | Hippocampus Volume | 0.027 | 0.005 | 0.05 | 0.016 | 0.28 | 0.82 |
| Processing Speed | Hippocampus Volume | 0.029 | 0.011 | 0.047 | 0.0014 | 0.26 | 0.85 |
| Language | Hippocampus Volume | 0.017 | -0.001 | 0.035 | 0.0617 | 0.22 | 0.77 |
| Visuospatial | Hippocampus Volume | -0.002 | -0.026 | 0.023 | 0.8991 | 0.09 | 0.43 |
| Global Composite | WMH Volume | -0.019 | -0.034 | -0.004 | 0.014 | 0.36 | 0.92 |
| Executive Functions | WMH Volume | -0.025 | -0.042 | -0.007 | 0.0052 | 0.29 | 0.86 |
| Episodic Memory | WMH Volume | -0.008 | -0.034 | 0.019 | 0.5705 | 0.25 | 0.82 |
| Processing Speed | WMH Volume | -0.024 | -0.045 | -0.004 | 0.022 | 0.26 | 0.85 |
| Language | WMH Volume | -0.013 | -0.035 | 0.008 | 0.2155 | 0.21 | 0.77 |
| Visuospatial | WMH Volume | 0.012 | -0.016 | 0.041 | 0.3962 | 0.09 | 0.43 |

**Supplementary Table 2 Results of the combined linear mixed effects models.**

| Outcome | Predictor | Estimate | CI Lower | CI Upper | *P* value | Marginal R2 | Conditional R2 |
| --- | --- | --- | --- | --- | --- | --- | --- |
| Global Composite | Aβ SUVR | -0.038 | -0.054 | -0.023 | <0.0001 | 0.43 | 0.92 |
| Global Composite | plasma p-tau217 | -0.022 | -0.039 | -0.006 | 0.0091 | 0.43 | 0.92 |
| Global Composite | plasma GFAP | -0.005 | -0.022 | 0.011 | 0.5324 | 0.43 | 0.92 |
| Global Composite | plasma NFL | -0.004 | -0.02 | 0.011 | 0.5961 | 0.43 | 0.92 |
| Global Composite | Hippocampus Volume | 0.017 | 0.006 | 0.028 | 0.002 | 0.43 | 0.92 |
| Global Composite | WMH Volume | -0.016 | -0.029 | -0.003 | 0.0178 | 0.43 | 0.92 |
| Executive Functions | Aβ SUVR | -0.038 | -0.057 | -0.018 | 0.0002 | 0.33 | 0.86 |
| Executive Functions | plasma p-tau217 | -0.019 | -0.041 | 0.003 | 0.0976 | 0.33 | 0.86 |
| Executive Functions | plasma GFAP | -0.012 | -0.033 | 0.01 | 0.2843 | 0.33 | 0.86 |
| Executive Functions | plasma NFL | -0.002 | -0.023 | 0.018 | 0.8134 | 0.33 | 0.86 |
| Executive Functions | Hippocampus Volume | 0.012 | -0.003 | 0.026 | 0.108 | 0.33 | 0.86 |
| Executive Functions | WMH Volume | -0.023 | -0.039 | -0.006 | 0.0072 | 0.33 | 0.86 |
| Episodic Memory | Aβ SUVR | -0.08 | -0.107 | -0.053 | <0.0001 | 0.36 | 0.83 |
| Episodic Memory | plasma p-tau217 | -0.021 | -0.051 | 0.01 | 0.1819 | 0.36 | 0.83 |
| Episodic Memory | plasma GFAP | -0.016 | -0.046 | 0.014 | 0.3078 | 0.36 | 0.83 |
| Episodic Memory | plasma NFL | -0.016 | -0.044 | 0.012 | 0.2636 | 0.36 | 0.83 |
| Episodic Memory | Hippocampus Volume | 0.022 | 0.002 | 0.041 | 0.0325 | 0.36 | 0.83 |
| Episodic Memory | WMH Volume | -0.002 | -0.026 | 0.021 | 0.8457 | 0.36 | 0.83 |
| Processing Speed | Aβ SUVR | -0.001 | -0.024 | 0.023 | 0.9618 | 0.29 | 0.85 |
| Processing Speed | plasma p-tau217 | -0.021 | -0.048 | 0.006 | 0.132 | 0.29 | 0.85 |
| Processing Speed | plasma GFAP | -0.007 | -0.034 | 0.02 | 0.6042 | 0.29 | 0.85 |
| Processing Speed | plasma NFL | -0.01 | -0.035 | 0.016 | 0.4457 | 0.29 | 0.85 |
| Processing Speed | Hippocampus Volume | 0.026 | 0.008 | 0.044 | 0.0044 | 0.29 | 0.85 |
| Processing Speed | WMH Volume | -0.021 | -0.042 | 0 | 0.0463 | 0.29 | 0.85 |
| Language | Aβ SUVR | -0.026 | -0.052 | 0 | 0.0476 | 0.25 | 0.77 |
| Language | plasma p-tau217 | -0.015 | -0.044 | 0.014 | 0.3019 | 0.25 | 0.77 |
| Language | plasma GFAP | 0.017 | -0.011 | 0.045 | 0.2382 | 0.25 | 0.77 |
| Language | plasma NFL | -0.016 | -0.042 | 0.011 | 0.2565 | 0.25 | 0.77 |
| Language | Hippocampus Volume | 0.014 | -0.005 | 0.032 | 0.1416 | 0.25 | 0.77 |
| Language | WMH Volume | -0.014 | -0.036 | 0.007 | 0.1965 | 0.25 | 0.77 |
| Visuospatial | Aβ SUVR | -0.01 | -0.045 | 0.025 | 0.566 | 0.11 | 0.44 |
| Visuospatial | plasma p-tau217 | -0.024 | -0.062 | 0.015 | 0.228 | 0.11 | 0.44 |
| Visuospatial | plasma GFAP | 0.007 | -0.031 | 0.045 | 0.7261 | 0.11 | 0.44 |
| Visuospatial | plasma NFL | -0.013 | -0.049 | 0.023 | 0.4703 | 0.11 | 0.44 |
| Visuospatial | Hippocampus Volume | -0.002 | -0.027 | 0.023 | 0.8892 | 0.11 | 0.44 |
| Visuospatial | WMH Volume | 0.012 | -0.018 | 0.041 | 0.4404 | 0.11 | 0.44 |

**Supplementary Table 3 Results of the WMH interaction linear mixed effects models.**

| Outcome | Predictor | Estimate | CI Lower | CI Upper | *P* value | Marginal R2 | Conditional R2 |
| --- | --- | --- | --- | --- | --- | --- | --- |
| Global Composite | Aβ SUVR | -0.018 | -0.027 | -0.009 | 0.0001 | 0.46 | 0.92 |
| Executive Functions | Aβ SUVR | -0.014 | -0.025 | -0.002 | 0.0195 | 0.33 | 0.86 |
| Episodic Memory | Aβ SUVR | -0.042 | -0.057 | -0.027 | <0.0001 | 0.47 | 0.82 |
| Processing Speed | Aβ SUVR | -0.006 | -0.02 | 0.009 | 0.4416 | 0.26 | 0.85 |
| Language | Aβ SUVR | -0.007 | -0.022 | 0.008 | 0.3855 | 0.23 | 0.77 |
| Visuospatial | Aβ SUVR | 0.026 | 0.006 | 0.046 | 0.0118 | 0.1 | 0.43 |
| Global Composite | plasma p-tau217 | -0.028 | -0.043 | -0.014 | 0.0001 | 0.41 | 0.92 |
| Executive Functions | plasma p-tau217 | -0.025 | -0.044 | -0.006 | 0.0106 | 0.31 | 0.86 |
| Episodic Memory | plasma p-tau217 | -0.043 | -0.069 | -0.017 | 0.0011 | 0.33 | 0.82 |
| Processing Speed | plasma p-tau217 | -0.028 | -0.05 | -0.006 | 0.0125 | 0.28 | 0.85 |
| Language | plasma p-tau217 | -0.036 | -0.06 | -0.012 | 0.0033 | 0.22 | 0.77 |
| Visuospatial | plasma p-tau217 | 0.017 | -0.014 | 0.048 | 0.2759 | 0.1 | 0.44 |
| Global Composite | plasma GFAP | -0.018 | -0.03 | -0.005 | 0.0059 | 0.43 | 0.92 |
| Executive Functions | plasma GFAP | -0.017 | -0.031 | -0.002 | 0.0275 | 0.32 | 0.86 |
| Episodic Memory | plasma GFAP | -0.033 | -0.055 | -0.011 | 0.0028 | 0.32 | 0.82 |
| Processing Speed | plasma GFAP | -0.019 | -0.037 | -0.001 | 0.0343 | 0.29 | 0.85 |
| Language | plasma GFAP | -0.015 | -0.034 | 0.004 | 0.1111 | 0.23 | 0.77 |
| Visuospatial | plasma GFAP | 0.045 | 0.02 | 0.069 | 0.0003 | 0.13 | 0.43 |
| Global Composite | plasma NFL | -0.004 | -0.017 | 0.009 | 0.5673 | 0.38 | 0.92 |
| Executive Functions | plasma NFL | 0.003 | -0.014 | 0.019 | 0.7553 | 0.31 | 0.86 |
| Episodic Memory | plasma NFL | -0.017 | -0.04 | 0.006 | 0.1553 | 0.26 | 0.82 |
| Processing Speed | plasma NFL | -0.013 | -0.032 | 0.005 | 0.159 | 0.28 | 0.85 |
| Language | plasma NFL | -0.026 | -0.046 | -0.006 | 0.0121 | 0.23 | 0.77 |
| Visuospatial | plasma NFL | 0.027 | 0.001 | 0.054 | 0.041 | 0.1 | 0.44 |
| Global Composite | Hippocampus Volume | 0.013 | 0.002 | 0.025 | 0.0236 | 0.41 | 0.92 |
| Executive Functions | Hippocampus Volume | 0.008 | -0.006 | 0.022 | 0.247 | 0.31 | 0.86 |
| Episodic Memory | Hippocampus Volume | 0.036 | 0.015 | 0.056 | 0.0007 | 0.34 | 0.82 |
| Processing Speed | Hippocampus Volume | -0.004 | -0.02 | 0.012 | 0.6313 | 0.26 | 0.85 |
| Language | Hippocampus Volume | -0.002 | -0.019 | 0.015 | 0.7943 | 0.23 | 0.77 |
| Visuospatial | Hippocampus Volume | -0.022 | -0.044 | 0.001 | 0.0638 | 0.09 | 0.43 |

**Supplementary Table 4 Results of the linear mixed effects model testing the interactive association between Fazekas Score, Amyloid SUVR, and time on episodic memory performance.**

| Predictor | Value | Std. Error | *P* value |
| --- | --- | --- | --- |
| (Intercept) | -0.088 | 0.192 | 0.645 |
| Years since Baseline | 0.014 | 0.031 | 0.639 |
| Amyloid SUVR | -0.014 | 0.275 | 0.959 |
| Fazekas (Score 1) | -0.167 | 0.19 | 0.383 |
| Fazekas (Score 2) | 0.08 | 0.209 | 0.701 |
| Fazekas (Score 3) | -0.046 | 0.252 | 0.857 |
| Age | -0.386 | 0.071 | <0.001 |
| Sex (male) | -0.338 | 0.133 | 0.012 |
| APOE4 | -0.009 | 0.146 | 0.949 |
| Education | 0.208 | 0.066 | 0.002 |
| Years since Baseline:Amyloid SUVR | -0.028 | 0.045 | 0.537 |
| Years since Baseline:Fazekas (Score 1) | -0.016 | 0.031 | 0.61 |
| Years since Baseline:Fazekas (Score 2) | -0.016 | 0.034 | 0.632 |
| Years since Baseline:Fazekas (Score 3) | -0.014 | 0.042 | 0.732 |
| Amyloid SUVR:Fazekas (Score 1) | 0.174 | 0.292 | 0.553 |
| Amyloid SUVR:Fazekas (Score 2) | -0.166 | 0.301 | 0.582 |
| Amyloid SUVR:Fazekas (Score 3) | -0.682 | 0.306 | 0.027 |
| Years since Baseline:Age | -0.029 | 0.012 | 0.019 |
| Years since Baseline:Sex (male) | 0.042 | 0.022 | 0.063 |
| Years since Baseline:APOE4 | -0.014 | 0.024 | 0.559 |
| Years since Baseline:Education | -0.002 | 0.011 | 0.835 |
| Years since Baseline:Amyloid SUVR:Fazekas (Score 1) | -0.041 | 0.048 | 0.391 |
| Years since Baseline:Amyloid SUVR:Fazekas (Score 2) | -0.074 | 0.05 | 0.143 |
| Years since Baseline:Amyloid SUVR:Fazekas (Score 3) | -0.122 | 0.051 | 0.018 |

**Supplementary Figure 1 Baseline associations between biomarkers and cognitive performance.** Estimates for hippocampus volume were reversed for a better comparison. Error bars represent 95% confidence intervals. All variables were standardized to allow direct comparison of effect sizes. All models were adjusted for age, sex, APOE ε4 status, education, and their interactions with time.

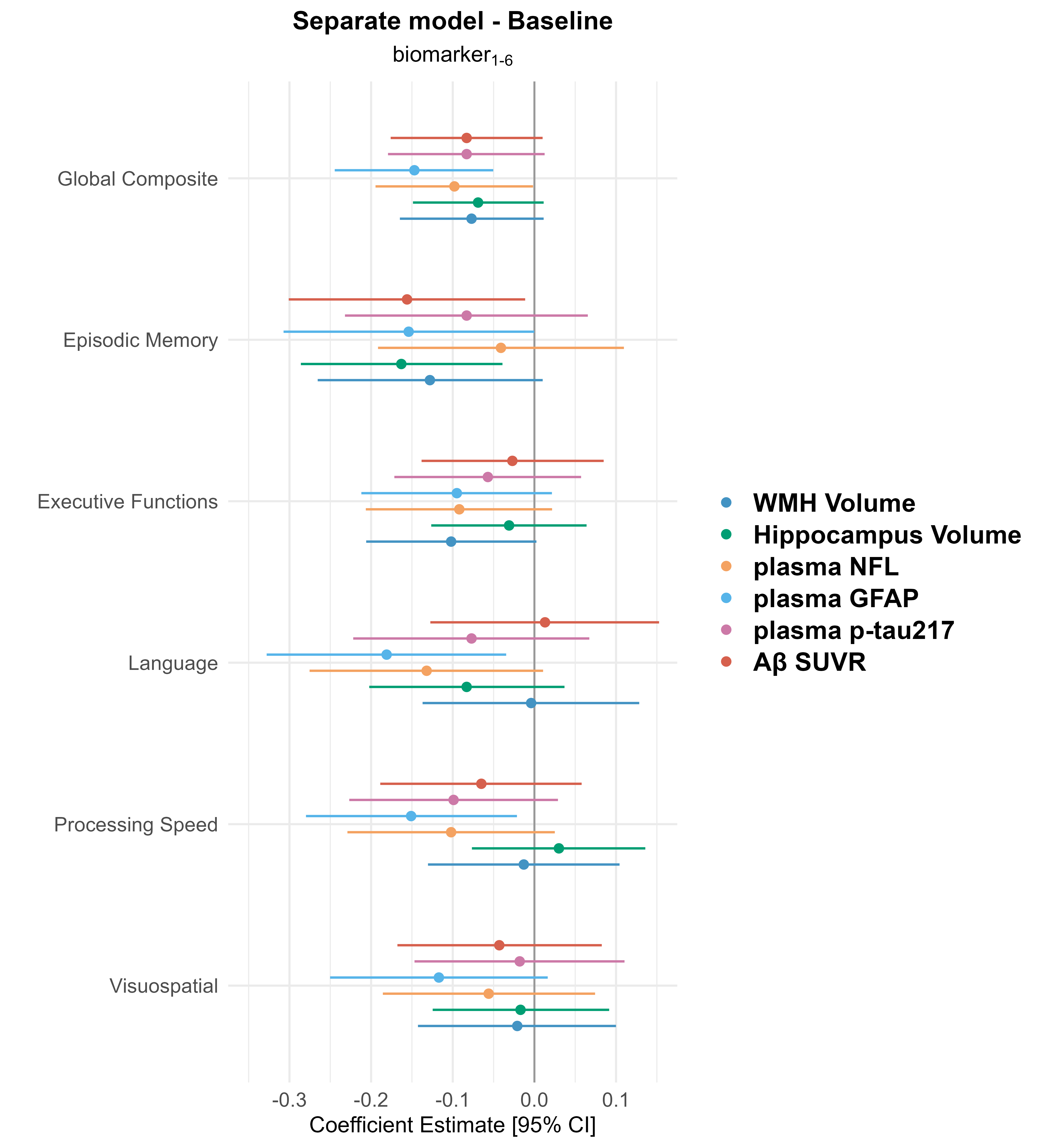

**Supplementary Figure 2 Interaction of WMH with amyloid, GFAP, and NfL on Visuospatial abilities visualized in low and high WMH groups.** Estimates in the plot are from separate linear-mixed effects models in group with low and high WMH volume.

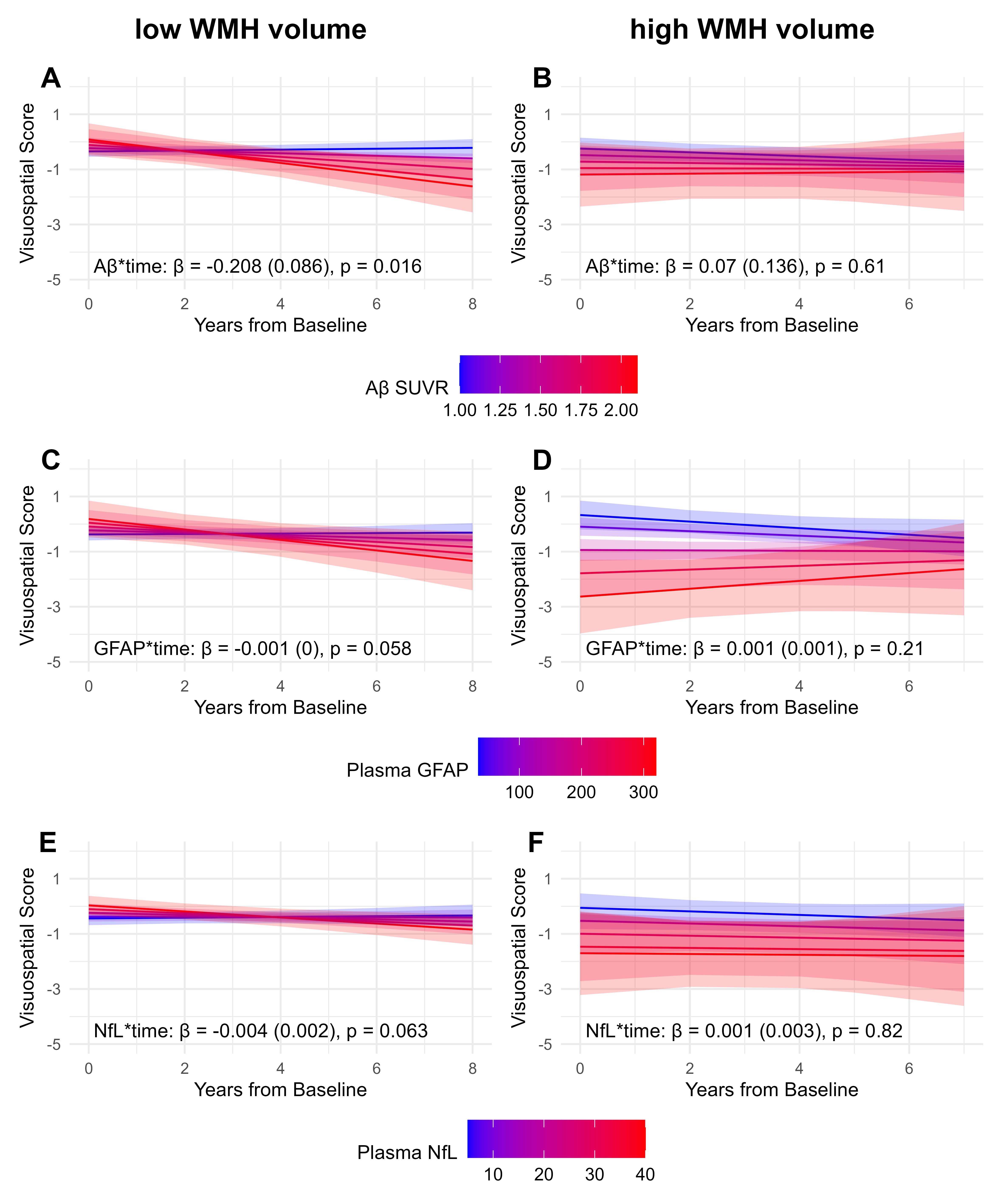

**Supplementary Figure 3 Results of the linear mixed-effects models adjusted for BMI and eGFR.** Estimates for hippocampus volume were reversed for a better comparison. Error bars represent 95% confidence intervals. All variables were standardized to allow direct comparison of effect sizes. In addition to BMI and eGFR, all models were adjusted for age, sex, APOE ε4 status, education, and their interactions with time.

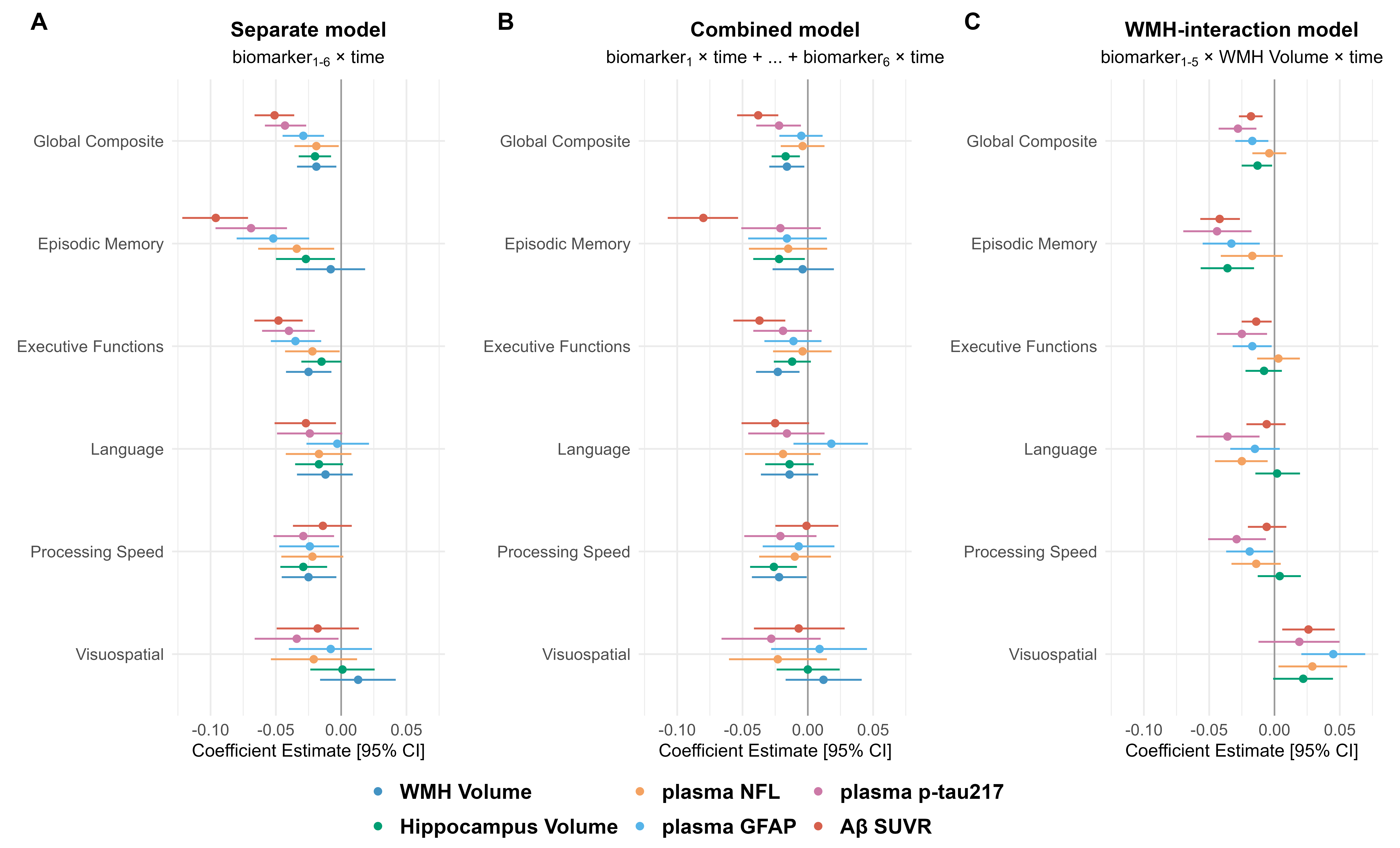

**Supplementary Figure 4 Results of the linear mixed-effects models adjusted for the presence or absence of hypertension.** Estimates for hippocampus volume were reversed for a better comparison. Error bars represent 95% confidence intervals. All variables were standardized to allow direct comparison of effect sizes. In addition to hypertension, all models were adjusted for age, sex, APOE ε4 status, education, and their interactions with time.

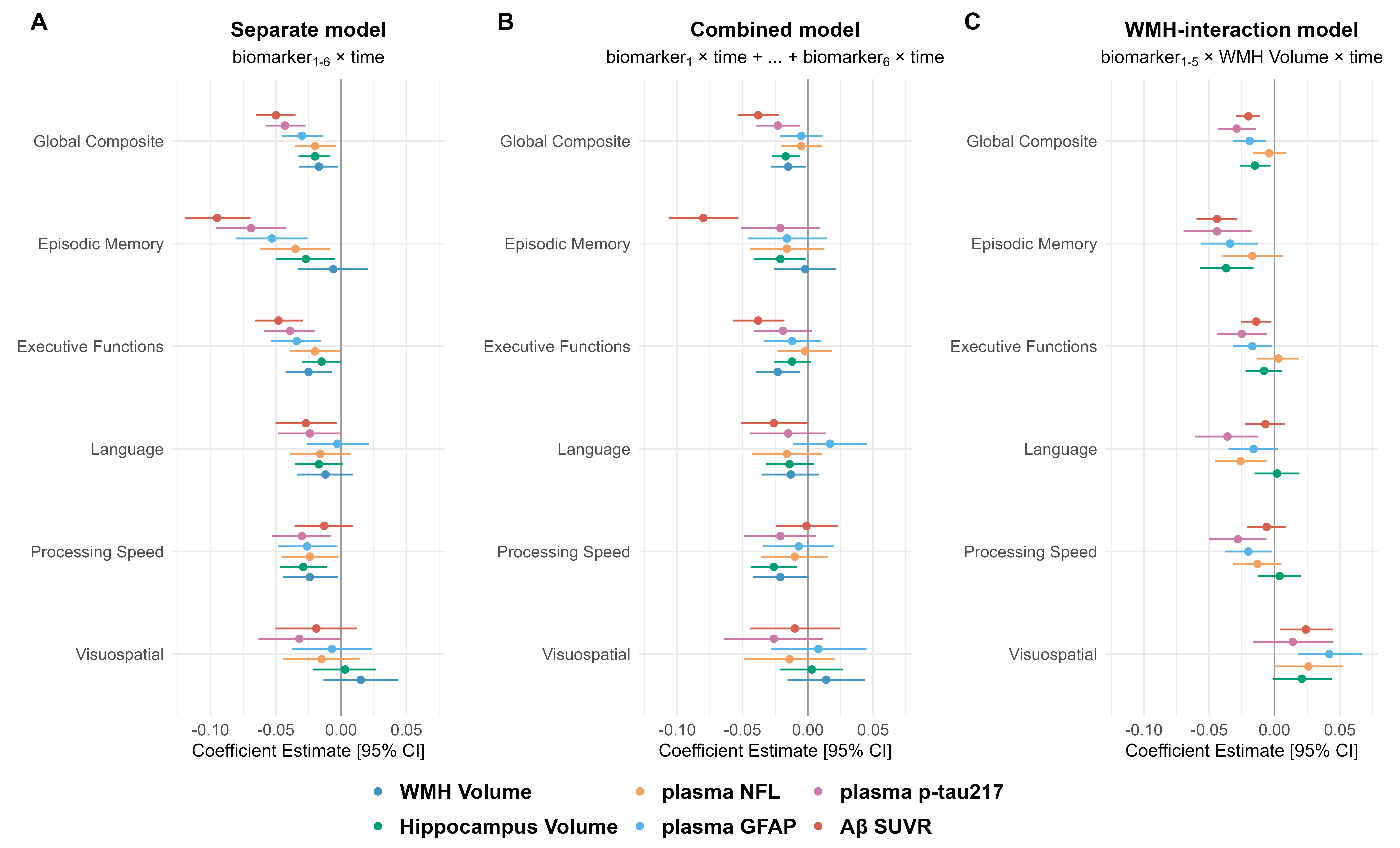

**Supplementary Figure 5 Results of the linear mixed-effects models when participants with a baseline CDR >0 were excluded.** Estimates for hippocampus volume were reversed for a better comparison. Error bars represent 95% confidence intervals. All variables were standardized to allow direct comparison of effect sizes. In addition to BMI and eGFR, all models were adjusted for age, sex, APOE ε4 status, education, and their interactions with time.

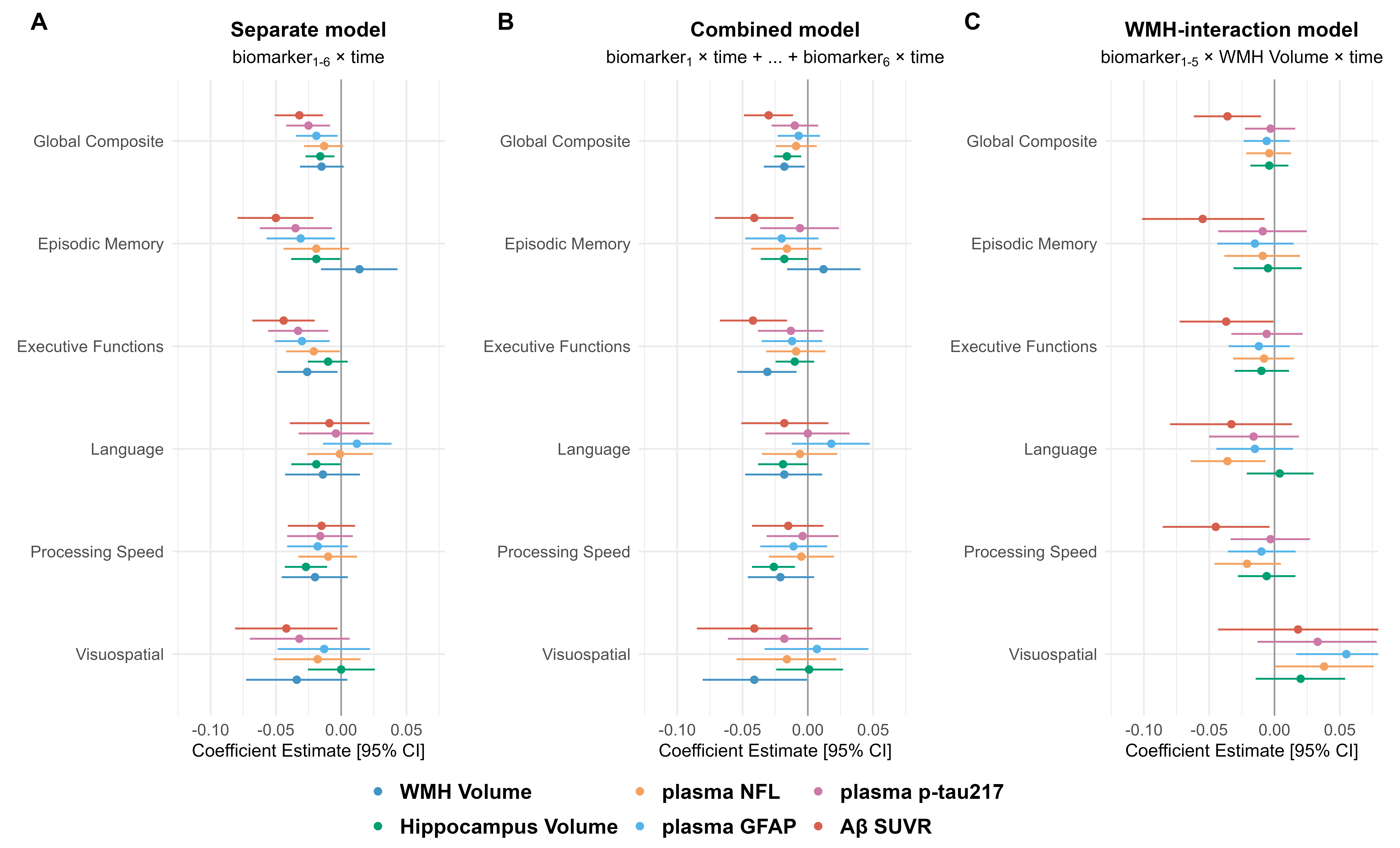

**Supplementary Figure 6 Final multigroup path analysis model with global cognition slope as the final outcome variable.** All paths were adjusted for age, sex, and APOE ε4 status. Paths to global cognition slope were additionally adjusted for education. All variables were standardized prior to model entry. Values in brackets indicate 95% confidence intervals, derived from 1000 bootstrap samples.

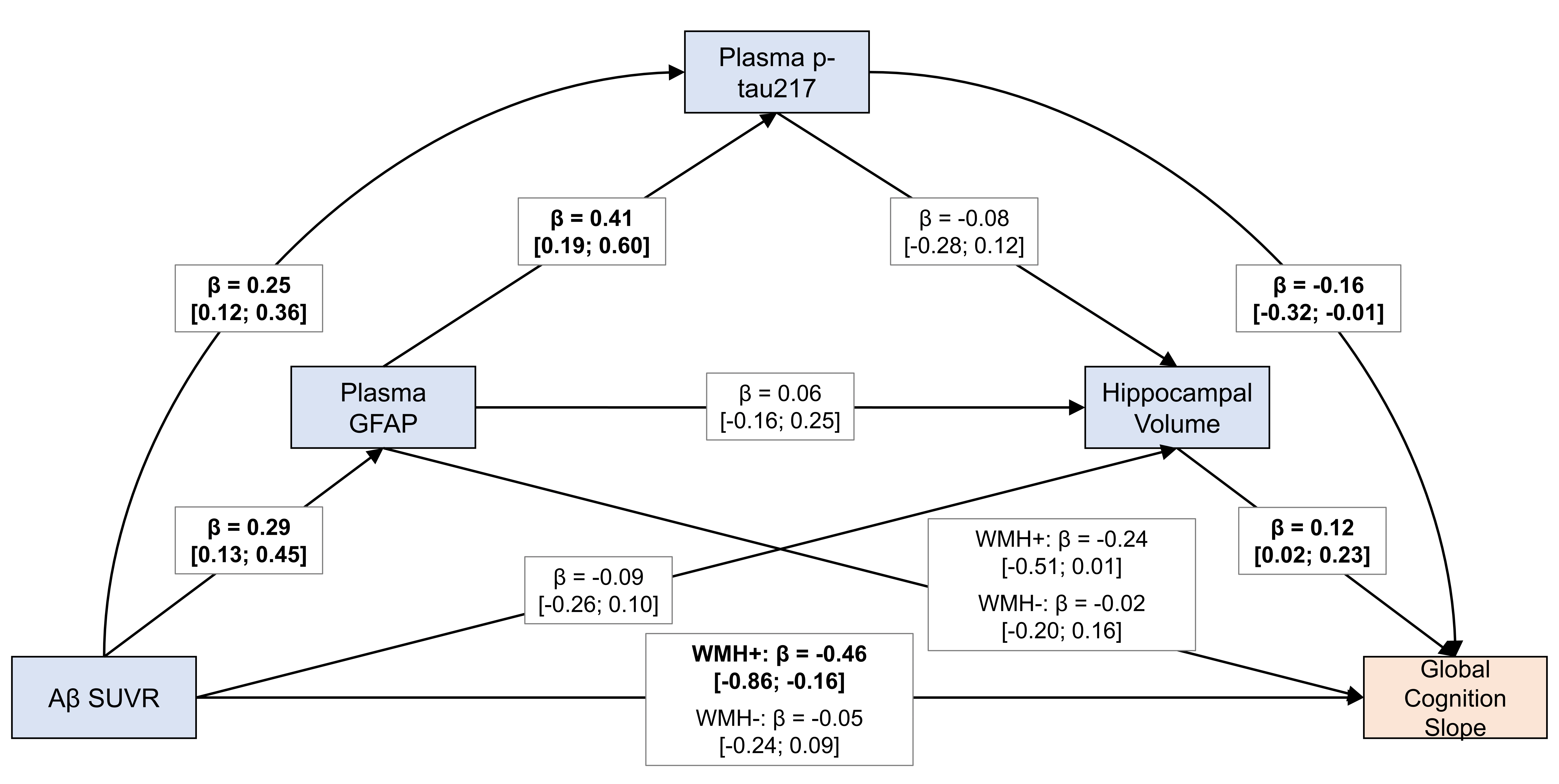

**Supplementary Figure 7 Distribution of total WMH percentage of TIV by Fazekas Score.** Boxplots showing the distribution of total WMH percentage across Fazekas score categories. Wilcoxon rank-sum tests were used to assess statistical differences between categories, with significance levels indicated on the plot. The horizontal red line indicates a WMH volume of 0.25% of the TIV, which was used to define elevated WMH in some of our previous analyses.

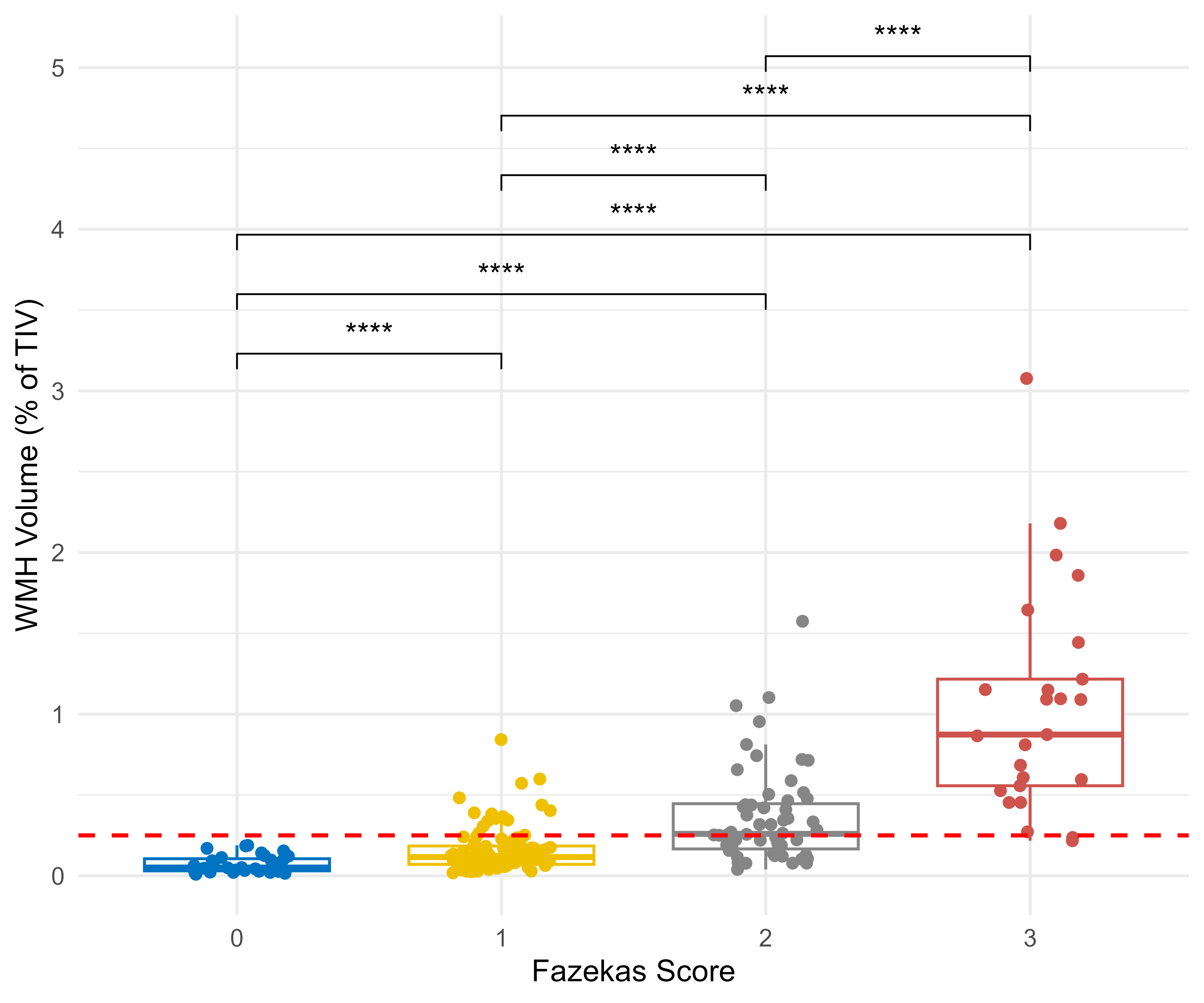
